# Knowledge, attitude and practice towards the novel corona virus among Bangladeshi people: Implications for mitigation measures

**DOI:** 10.1101/2020.05.05.20091181

**Authors:** Alak Paul, Dwaipayan Sikdar, Mohammad Mosharraf Hossain, Md Robed Amin, Farah Deeba, Janardan Mahanta, Md. Akib Jabed, Mohammad Mohaiminul Islam, Sharifa Jahan Noon, Tapan Kumar Nath

## Abstract

The current novel coronavirus (nCoV) outbreak, COVID-19, was first reported in December 2019 in Wuhan, China has spread all over the world causing startling loss of lives, stalling the global economy and disrupting the social life. One of the challenges to contain the COVID-19 is making people adopt personal hygiene, social distancing and self-quarantine practices which are all related to knowledge, attitude and practice (KAP) of the people in respective countries. Bangladesh, the most densely populated countries with a fast-growing economy and moderate literacy rate, has shown many hiccups in its efforts to implement COVID-19 policies. Understanding KAP may give the policy makers to make informed decisions. Hence, this study aimed to make a quick assessment of KAP of people in relation to COVID-19 in Bangladesh. An online survey using a pre-tested questionnaire conducted in late March 2020 attained 1837 responses across Bangladesh. However, 1589 completed responses were included in statistical analysis to calculate KAP scores, their interrelations with socio-demographic variables. The overall KAP is poor with only 10% of the respondents showed good knowledge with parallel attitudes and practices. Socio-demographic factors have strong bearings on the KAP scores. Significantly higher KAP score is evident in female over male respondents, among aged (45 years and above) over younger respondents and among retired and homemakers above students and public service holders. The study indicated a panic fuelled by poor understanding of COVID-19 associated facts and the need for the government to ensure more granular and targeted awareness campaigns in a transparent and factual manner to gain public confidence and arrest more meaningful public participation in mitigation measures. The study provides a baseline of KAP among people in Bangladesh on COVID-19.

## Introduction

Alike all new years, 2020 started with new hopes and resolutions only to be shattered by the rapidly unfolding onslaught of corona virus disease 2019 (COVID-19) from the novel coronavirus (nCOV, later named as SARS-CoV-2) that originated from unknown source in Wuhan, China [1-3]. Unlike previous corona outbreaks [4], this highly contagious [5–9] zoonotic virus from yet to confirm animal origin [10-11], has gone from a local flue related severe acute respiratory syndrome [4,8,12,13] to a pandemic threatening lives of millions within a span of few weeks. COVID-19 has thrown global public health into a turmoilby severely straining the healthcare system of the world irrespective of countries. The rapid movement of epicentre moved from China to Iran and then through Europe to USA over a span of nine weeks awed us not only about COVID-19 but also the looming future infectious diseases [14]. As it spreads through social contacts [15,16], billions of people have already been forced into lockdown to minimize the rate of spread that infected subsequent hundred thousand people in a couple of months putting them through economic and psychological hardship [4]. The lockdown has to be adopted since researchers need time to come up with a vaccine or effective treatment as in preceding pandemics including SARS and MARS [4,17-18]. No imminent solution for COVID-19 is in view in the immediate future [19].

The healthcare system of the first-world has failed to tackle the challenge to provide medical care for the rapidly increasing number of infected patients, let alone the developing or underdeveloped nations [20-21]. In the majority of the cases, the leadership in different countries along with the bureaucracy seemed indecisive, inefficient, unpreparedand at awe to contain the contagion. Eventually, for the first time in global history, the active participation of every single human being on earth - in the form of testing, isolation, contact tracing, social distancing, stay at home, self-quarantine and personal hygiene and use of personal protective equipment likes masks and gloves – has become critical to the containment of COVID-19 to allow the hospitals a lax for not being overwhelmed and the researchers some time to come up with a remedy [22-23]. In this global drill, everyone has to sacrifice - their autonomy, health, job, business, recreation, education, and whatnot. However, ensuring voluntary participation of people in the drill has posed varying challenges in different countries due to varying levels of Knowledge, Attitudes, and Practices (KAP) of the people. Accordingly, design and success of all anti-contagion initiatives depends on macro- and micro-level understanding of KAP in respective countries and within each country.

In Bangladesh, like the SAARC region, the COVID-19 situation seems grave though it poses otherwise initially [24,25] mainly due to imported cases from expatriates [26]. Following its first positive COVID-19 case on March8, 2020 [27], the country has stopped its educational institutions since March 17, saw its first COVID-19 death on 18 March [28] declared the closure of everything on a halt from March 26 and involved police and army to strengthen the implementation of lockdown as the country is a densely populated, depends on labour-oriented industries [29] and a vast majority thrives on daily earning through informal occupations [30]. However, the lack of coordination of response to the threat is evident [27] which indicates design and implementation of these initiatives based on poor understanding of the KAPs of socio-demographic groups. Hence, this research aimed to quickly assess the Knowledge, Attitudes, and Practices of people in Bangladesh on nCOV by taking netizens as a representative sample with the hope that the outcome will assist the authorities and other stakeholders to improve the planning and execution of different measures and will provide a reference for countries with similar socio-demographic.

## Methods

### Conceptual framework

This study followed KAP approach because this is a representative tool used for a specific population to collect information on what is known, believed, and done in relation to a specific field, for example health [31]. Historically, the KAP model was developed for family planning and population studies in the 1950s, with its purpose being to measure the extent to which any clear opposition to the notion and organization of family planning existed among different populations, so specific family planning practices could be used for different program purposes worldwide [31]. KAP surveys are now the most widely used studies for uncovering societal context in public health research [32-34]. These surveys are easy to design, data output is quantifiable, interpretation is robust and their utility is generalizable for context specific problems [35]. Information generated through KAP studies assist to develop strategies with a focus to improve the behavioral and attitudinal changes driven by the level of knowledge and perceptions towards preventive practices [36]. In a recent KAP study conducted in China, Zhong et al. [37] reported that to facilitate outbreak management of COVID-19, there is an urgent need to understand the public’s awareness of COVID-19. They asserted that success against COVID-19 requires peoples’ adherence to control measures which is largely affected by their knowledge, attitudes and practices.

### Instruments and participants

This KAP study was conducted across Bangladesh using an online survey. Because of contagious nature of COVID-19, we avoided physical interviews. Following Zhong et al. [37] who studied KAP in COVID-19 infected areas in China, and braining storming among authors of this paper, we prepared a structured questionnaire having 50 multiple choice questions. This was tested in a pilot study with 10 participants. Based on feedbacks from pilot study, we revised the questionnaire and finalized with 40 questions (S1 Table). The questionnaire had four parts-A) Basic information of participants (5 questions), B) Knowledge on Covid-19 (16 questions), C) Attitudes (10 questions) and D) Practice (9 questions). Using office 365, a form was created and the link of the same was shared through Facebook and email with brief introduction and objectives of the survey. Respondents were requested repeatedly to share the form widely to collect a snowball sample of representative of Bangladeshi population aged 18 and above. The survey anticipated responses from participants having university level education as the questions were written in English. Participation was voluntary, anonymous and could withdraw from survey at any time. They provided informed consent prior to complete the survey. The form was posted on 22 March 2020 at 22:00 and the survey was closed on 28 March 2020 at 00:15.

In order to contextualize respondents’ views with the public sectors’ preparedness in COVID-19 mitigation-relevant policy documents, press releases and newspaper reports were reviewed, synthesized and described following content analysis [50]. The ethical review committee of Dhaka Medical College, Bangladesh has approved this survey (Memo No. ERC-DMC/ECC/2020/88).

### Data cleaning

Respondents input their opinions and information by using the shared online survey form. Responses were automatically stored in Microsoft Excel. A total of 1837 respondents participated in this study. However, some of the respondents did not fully complete the survey questionnaire and those were discarded from the survey leaving 1589 complete responses.

### Scoring scheme

Respective scores of knowledge, attitude and practice for each respondent were obtained from their responses respectively in 13 knowledge questions, 10 attitude questions and nine practice questions. Percentage of correct answers in knowledge, attitude and practice questions yielded the scores of respective categories. A cut point of 80% correct answers was used for all categories to differentiate between *good* and *poor* knowledge, attitude and practices.

### Software

The online survey was conducted by distributing the KAP questionnaire as a Microsoft Office form through Facebook and emails. After importing the online survey results through MS Excel, R programming version 3.5.2 was used for raw data handling and for statistical analysis. Some statistical analyseses were performed on SPSS (Statistical Package for the Social Sciences) version 16.

### Statistical analysis

Based on the scores of different knowledge, attitude and practice variables, the mean difference between/among the categories of different socio-demographic characteristics were compared with independent samples t-test (for two categories of variable), and one-way analysis of variance (ANOVA)/F-test (for more than two categories of variable). Post hoc analyses were done for significant variable. Also check the associations between different knowledge, attitude and practice variables with different socio-demographic variables were shown using Chi square test. Logistic regressions were being run about the variables that were significant in bivariate analyses/Chi square test.

## Results and discussion

We have described socio-demographic information of respondents in the beginning followed by overall KAP scores, assessment of KAP responses, and relationship among attitude, practices and knowledge. This is the first study of this kind in Bangladesh, to the best of our knowledge, that focused on KAP related to COVID-19 and such studies from elsewhere is also quite limited. Therefore, discussion of results draws evidences from media reports.

### Socio-demographic characteristics of the respondents

A total of 1837 participants responded to the online questionnaire reached mainly through Facebook and emails, out of which 1589 questionnaires contained complete responses and were included into this analysis discarding the rest 248 responses because of missing data. The geographical distribution of respondents is shown in S1 Fig. The characteristics of study participants according to study site are summarized in Table 1. Among the respondents 961 (60.48%) were *males* and 628 (39.52%) were *females*. The distribution of respondents in terms of age groups was *‘18-25’* 739 (46.51%), *‘26-35’* 533 (33.54%), *‘36-45’* 184 (11.58%), *‘46-65’* 124 (7.80%), *‘above 65’* 9 (0.57%) which indicated that online survey reaches younger population disproportionately over the elders. Respondents educational status also exhibited the influence of online access as the makeup was *‘Secondary & Below’* (4.22%), *‘University’* (95.78%) as does their professional status which was made up of *‘Government Staff’* (4.59%), *‘Home* (2.71%), *‘Professional’* (40.28%), *‘Retired’* (0.69%), *‘Student’* (44.49%), and *‘Unemployed’* (7.24%).

**Table 1.** Socio-demographic profile of the respondents.

| Categories | Groups | Frequency | Percentage |
| --- | --- | --- | --- |
| **Gender** | Male | 961 | 60.5 |
|  | Female | 628 | 39.5 |
| **Age group** | 18-25 | 739 | 46.5 |
|  | 26-35 | 533 | 33.5 |
|  | 36-45 | 184 | 11.6 |
|  | 46-65 | 124 | 7.8 |
|  | Above 65 | 9 | 0.6 |
| **Education** | Secondary & Below | 67 | 4.2 |
|  | University | 1522 | 95.8 |
| **Occupation** | Government staff | 73 | 4.6 |
|  | Homemakers | 43 | 2.7 |
|  | Professionals | 640 | 40.3 |
|  | Retired | 11 | 0.7 |
|  | Student | 707 | 44.5 |
|  | Unemployed | 115 | 7.2 |

### KAP scores of Bangladeshi netizens on COVID-19

Overall, the people of Bangladesh demonstrated poor knowledge scores with a mean score of (8.56±1.63, on a scale of 13.0). As we see in Table 2, there is no statistically significant difference in knowledge score between male (8.57±1.70) and female (8.55±1.51) – a good outcome due to a wide range of educational support for females in the country. Surprisingly, the difference in knowledge score is also insignificant between educational groups. In the contrary, age and occupation show statistically significant (p<0.01) bearing on knowledge score. Among age groups, elders are more knowledgeable on COVID-19 than younger. The highest knowledge score (9.11±1.58) is in *‘46-65’* age group followed by (9.00 ±1.58) in ‘*Above 65* age group. The lowest score (8.30±1.70) is in *‘18-25’* age group while *‘26-35’* and *‘36-45’* age groups show similar knowledge scores (8.73±1.47 and 8.74±1.65, respectively). Retired people shows the highest knowledge score (9.27±1.62) on COVID-19 followed by government staffs (8.82±1.45) and professionals (8.80±1.45). Contrary to expectation, students showed the lowest knowledge score (8.33±1.66) subsequent to the homemakers. Comparatively lower knowledge scores among unemployed (8.87±2.01) and homemakers (8.37±1.60) and unemployed (8.53±1.87). Preceding observations indicated an overall poor knowledge score across socio-demographics as, being a recent phenomenon, people are unfamiliar with COVID-19, and awareness campaigns falls short in reaching all groups equally.

**Table 2.** Socio-demographic distribution of the respondents and their knowledge scores.

| Demographic variables | | Knowledge score in 13.00 ($\bar{x} \pm s$) | $t / F$ test | P-Value | Attitude score in 9.00 ($\bar{x} \pm s$) | $t / F$ test | P-Value | Practice score in 9.00 ($\bar{x} \pm s$) | $t / F$ test | P-Value |
| --- | --- | --- | --- | --- | --- | --- | --- | --- | --- | --- |
| Gender | Male | $8.57 \pm 1.70$ | 0.346 | 0.700 | $7.12 \pm 1.15$ | -4.61 | 0.000*** | $5.73 \pm 1.39$ | -6.01 | 0.000*** |
| | Female | $8.55 \pm 1.51$ | | | **$7.38 \pm 1.03$** | | | **$6.15 \pm 1.26$** | | |
| Age Group (years) | 18-25 | $8.30 \pm 1.70$ | 10.39 | 0.000*** | $7.16 \pm 1.13$ | 1.22 | 0.300 | $5.91 \pm 1.36$ | 2.878 | 0.022** |
| | 26-35 | $8.73 \pm 1.47$ | | | $7.30 \pm 1.08$ | | | **$5.94 \pm 1.32$** | | |
| | 36-45 | $8.74 \pm 1.65$ | | | $7.21 \pm 1.16$ | | | $5.89 \pm 1.45$ | | |
| | 46-65 | **$9.11 \pm 1.58$** | | | $7.26 \pm 1.10$ | | | $5.54 \pm 1.33$ | | |
| | Above 65 | **$9.00 \pm 1.58$** | | | $7.44 \pm 0.73$ | | | **$6.56 \pm 1.01$** | | |
| Education | $\leq 12$ years | $8.43 \pm 1.66$ | -0.67 | 0.502 | $6.97 \pm 1.27$ | -1.925 | 0.054 | $5.63 \pm 1.24$ | -1.659 | 0.097 |
| | $> 12$ years | $8.57 \pm 1.63$ | | | $7.24 \pm 1.10$ | | | $5.91 \pm 1.36$ | | |
| Occupation | Govt. Staff | **$8.82 \pm 1.45$** | 6.63 | 0.000*** | $7.08 \pm 1.22$ | 3.86 | 0.002*** | $5.40 \pm 1.36$ | 3.72 | 0.002*** |
| | Homemakers | $8.37 \pm 1.60$ | | | **$7.42 \pm 0.82$** | | | **$6.28 \pm 1.28$** | | |
| | Professional | $8.80 \pm 1.54$ | | | $7.35 \pm 1.04$ | | | $5.86 \pm 1.37$ | | |
|  | Retired | **$9.27 \pm 1.62$** |  |  | **$7.55 \pm 0.69$** |  |  | **$6.45 \pm 1.29$** |  |  |
| | Student | $8.33 \pm 1.66$ | | | $7.13 \pm 1.17$ | | | $5.92 \pm 1.35$ | | |
| | Unemployed | $8.53 \pm 1.87$ | | | $7.08 \pm 1.16$ | | | $6.10 \pm 1.26$ | | |

\*\*\*Significant at 0.01 level \*\*Significant at 0.05 level (2-tailed).

The mean attitude score (7.23±1.11) on the scale of 9.00 indicated the desired attitude towards COVID-19 among Bangladeshi people (Table 2). On the other hand, the attitude scores varied significantly (p<0.01) between gender and among occupation groups. The attitude score for female (7.38±1.03) is higher than that for male (7.12±1. 15) though both have comparable knowledge scores. This observation is a positive one as females are more responsible in maintaining family hygiene and teach the children. The good knowledge, attitude and practice of female over male is supported by their lower mortality rate in this diseases, although female are globally more prone to this disease than male [32,42]. While the highest attitude score of the *‘Retired’* (7.55±0.69) was a reflection of their highest knowledge score; for other professional groups, knowledge scores mismatched their attitude scores. Surprisingly, the attitude scores of *‘Government staff’* (7.08±1.22) was the lowest with that of the *‘Student’* (7.08±1.16). *‘Homemakers’* and Professionals showed better attitude scores than *‘Unemployed’, ‘Government Staff’*, and *‘Students’*.

In stark contrast to knowledge and attitude scores, the mean practice score was poor (5.90±1.36 on the scale of 9.00) across all socio-socio-demographic groups (Table 2). The low practice score strongly indicates the gap in the translation of knowledge and attitudes into practices which is worrisome but expected in a developing country with median literacy and daily-earner based economy. Also, knowledge scores significantly varied (p<0.01) between genders, among age and occupational groups. Females, eldest and retired people with respective practice scores of 6.15±1.26, 6.56± 1.01 and 6.45±1.29 exhibited better translation of knowledge and attitudes into practice. Poor knowledge, attitude and practices among the students and the public service professionals is concerning since the young people constitutes a substantial percentage of the population while the public service professionals execute the public policies and mitigation responses. Hence, special preference should be given to enrich the knowledge of these two groups while focusing on improving their attitudes and practices. Overall, statistically significant and positive linear correlations is evident between knowledge and practice (r=0.093, p<0.01), knowledge and attitude (r=0.254, p<0.01) and attitude and practice (r=0.128, p<0.01) as shown in S2 Table. The observed differences may be due to disproportionate exposure of different groups to media, information gathering network among other factors. For example, while students and young adults are more engaged in social media, the elderly, homemakers have more time to spend on television and gather better information and convert it into better. It indicates avenues of improvement in an awareness campaign to target the age groups and professional groups through appropriate media.

### Assessment of KAP responses

As we can see from the frequency distribution of respondents on KAP questions (Tables 3-5), 54.8% of the respondents have a factual concept about COVID-19 and identify it as a *deadly disease, curable and low mortality rate i.e*., almost half of the respondents has the wrong notion about the disease and 36.2% of them have identified COVID-19 as *a deadly disease with the certainty of death*. A staggering 82.8% of the respondents showed the wrong conception about the cause of the emergence of COVID-19. These observations indicate the effect of misinformation from the internet and media in their understanding of the cause of the emergence of COVID-19. Also, disagreement among the respondents regarding the risk of elderly having comorbid diseases is indicative of an inadequate understanding of the outcome of COVID-19. Similarly, the concept of contagiousness of nCOV is unclear to a handsome one-fifth of the respondents. About 50% of the respondents considered the use of surgical masks may be effective in preventing infection while 25% percept that masks are inadequate and the rest demonstrated confusion. It is linked to the indecision of policymakers and mixed messages regarding masks. These results commensurate with observations from instances of harsh treatment of people who are wither diagnosed with COVID-19 or showed symptoms [36,51]. The respondents demonstrated sound knowledge on the symptoms of COVID (~99%), need for every person to adopt preventive measures (~90%), the duration of quarantine (95%), the means of reducing the spread of COVID (98%) and understanding about the treatment (~98%) which indicates the outcome of related contents in the awareness campaigns. The concept of quarantine was satisfactory in 86% of the respondents while one-tenth of them wrongly considered staying at home with family members equates quarantine. However, the concept of the situation in which someone needs to go for quarantine was quite shallow as opinions varied markedly. Respondents’ knowledge on the risk of the spread of nCOV in Bangladesh compared to other countries is alarming as the majority (75%) choose the wrong options while eerily almost all of them (99%) are wrong in selecting the priority measures that government needs to adopt to stop the spread. They also demonstrated a poor understanding of factors associated with the spread of nCOV as almost half of them (47%) couldn’t choose the right options. All these observations are explained by the newspaper reports [24,40] related to peoples’ behaviour contradicting the measures needed to keep it under control and overwhelming despises on measures taken by the government. It indicates the need to better communicate the information to make people educated so that they can better grasp the communications from the government and respond positively by conforming to the right attitudes and practices.

**Table 3.** Frequency distribution of responses on knowledge questions.

| Knowledge | No. | % |
| --- | --- | --- |
| **Perception about COVID-19** |  |  |
| A curse from the God | 122 | 7.68 |
| A deadly disease with certainty of death | 575 | 36.19 |
| **A deadly disease, curable and low mortality rate** | **870** | **54.75** |
| A rumor which is being spread through public or media | 22 | 1.38 |
| **COVID-19 was emerged due to following reasons. Multiple answer is allowed.** |  |  |
| **Right answers** | **273** | **17.18** |
| Wrong answers | 1316 | 82.82 |
| **The main clinical symptoms of covid-19 are fever, fatigue, dry cough, and breathing difficulty** |  |  |
| May be | 176 | 11.08 |
| No | 4 | 0.25 |
| **Yes** | **1409** | **88.67** |
| **Currently there is no effective cure for covid-2019, but early symptomatic and supportive treatment can help most patients recover from the infection** |  |  |
| May be | 293 | 18.44 |
| No | 32 | 2.01 |
| **Yes** | **1264** | **79.55** |
| **Only elderly people having chronic illnesses and other health complications are more likely to be seriously affected** |  |  |
| May be | 349 | 21.96 |
| No | 425 | 26.75 |
| **Yes** | **815** | **51.29** |
| **Persons with covid-2019 having no fever cannot infect to others.** |  |  |
| May be | 171 | 10.76 |
| **No** | **1262** | **79.42** |
| Yes | 156 | 9.82 |
| **The covid-19 spreads via respiratory droplets (from coughing, sneezing) of infected people.** |  |  |
| May be | 82 | 5.16 |
| No | 25 | 1.57 |
| **Yes** | **1482** | **93.27** |
| **Ordinary residents can wear general medical masks to prevent covid-19 infection.** |  |  |
| May be | 397 | 24.98 |
| **No** | **392** | **24.67** |
| Yes | 800 | 50.35 |
| **It is not necessary for children and young adults to take measures to prevent the infection by the covid-19.** |  |  |
| May be | 31 | 1.95 |
| **No** | **1425** | **89.68** |
| Yes | 133 | 8.37 |
| **What do you understand by quarantine?** |  |  |
| No clear understanding | 19 | 1.20 |
| **Stay at a separate room, and no contact with family members** | **1368** | **86.09** |
| Stay at home and can go outside | 23 | 1.45 |
| Stay at home with family members | 180 | 11.33 |
| **When should we go for quarantine? Multiple answer is allowed.** |  |  |
| **Right answer** | **102** | **6.42** |
| Wrong answer | 1487 | 93.58 |
| **Quarantine period** |  |  |
| **Right answer** | **1516** | **95.41** |
| Wrong answer | 73 | 4.59 |
| **Isolation and treatment of covid-19 infected people are effective ways to reduce the spread of the virus.** |  |  |
| May be | 130 | 8.18 |
| No | 28 | 1.76 |
| **Yes** | **1431** | **90.06** |
| **Comparing with other affected nations, what is the possibility of COVID-19 spread in Bangladesh? (Multiple answers).** |  |  |
| **Right answers** | **399** | **25.11** |
| Wrong answers | 1190 | 74.89 |
| **What could be the possible reasons of COVID-19 spread in Bangladesh if it happens? (Multiple answers)** |  |  |
| **Right answers** | **842** | **52.99** |
| Wrong answers | 747 | 47.01 |
| **What should be the priority actions for government to control the spread of covid-19? (Multiple answers)** |  |  |
| **Right answer** | **9** | **0.57** |
| Wrong answers | 1580 | 99.43 |

**Table 4.** Frequency distribution of responses on attitude questions.

| Attitude |  |  |
| --- | --- | --- |
|  | No. | % |
| *Do you like to stay at home for certain period (14 days) to prevent covid-19 spread if government will order so?* |  |  |
| Yes | 1512 | 95.15 |
| No | 18 | 1.13 |
| Not possible due to work | 59 | 3.71 |
| *Do you think that social distancing (e.g. stay 1-2 m apart, avoid crowds, etc.) can prevent covid-19 spread?* |  |  |
| May be | 300 | 18.88 |
| No | 74 | 4.66 |
| Yes | 1215 | 76.46 |
| *Do you agree that we should cancel business/recreational trips at this time?* |  |  |
| May be | 33 | 2.08 |
| No | 21 | 1.32 |
| Yes | 1535 | 96.60 |
| *Do you believe that working from home can help control covid-19?* |  |  |
| May be | 162 | 10.20 |
| No | 25 | 1.57 |
| Yes | 1402 | 88.23 |
| *Do you agree that government has taken sufficient preventive measures to prevent the spread of covid-19?* |  |  |
| No | 823 | 51.79 |
| Not Enough | 631 | 39.71 |
| Yes | 135 | 8.50 |
| *Do you agree that government should take preventive measures when covid-19 was first reported in China?* |  |  |
| May be | 92 | 5.79 |
| No | 222 | 13.97 |
| Yes | 1275 | 80.24 |
| *Do you think covid-19 can cause massive fatality in Bangladesh?* |  |  |
| May be | 295 | 18.57 |
| No | 59 | 3.71 |
| Yes | 1235 | 77.72 |
| *Do you believe that COVID-19 will not be epidemic in Bangladesh due to following reasons? (Multiple answer)* |  |  |
| Right answers | 1081 | 68.03 |
| Wrong answers | 508 | 31.97 |
| *Do you agree that our health support providers (e.g. doctors, nurses, support staff) are under serious threat when they treat infected people?* |  |  |
| May be | 102 | 6.42 |
| No | 89 | 5.60 |
| Yes | 1398 | 87.98 |
| *Do you think that government has ensured enough protective measures for health support providers?* |  |  |
| May be | 119 | 7.49 |
| No | 1396 | 87.85 |
| Yes | 74 | 4.66 |

**Table 5.** Frequency distribution of responses on practices questions.

| Practices | No. | % |
| --- | --- | --- |
| *Do you go to crowded areas now-a-days?* |  |  |
| Every day | 14 | 0.88 |
| **No** | **1279** | **80.49** |
| Sometimes | 268 | 16.87 |
| Yes | 28 | 1.76 |
| *Do you allow your children to go for outdoor activities?* |  |  |
| **No** | **1498** | **94.27** |
| Sometimes | 78 | 4.91 |
| Yes | 13 | 0.82 |
| *Do you and family members use mask when go outside?* |  |  |
| **No** | **90** | **5.66** |
| Sometimes | 240 | 15.10 |
| Yes | 1259 | 79.23 |
| *Have you started working from home in last few weeks due to outbreak of covid-19?* |  |  |
| No | 420 | 26.43 |
| Sometimes | 229 | 14.41 |
| **Yes** | **940** | **59.16** |
| *How would you rate the awareness level of the people living around you regarding covid-19?* |  |  |
| Awareness level is on rise | 265 | 16.68 |
| **Little awareness has grown so far** | **724** | **45.56** |
| No precautionary measures undertaken at all | 176 | 11.07 |
| People around me are highly aware and careful | 70 | 4.41 |
| Some precautionary measures have been taken | 354 | 22.28 |
| *How would you rate the medical facilities in Bangladesh to handle covid-19?* |  |  |
| Gradual advancement in health care is noticeable to deal with covid-19 | 77 | 4.85 |
| Health facilities are available for limited number of people | 381 | 23.98 |
| Medical facilities are highly appreciable, and it can prevent the spread of covid-19 | 11 | 0.69 |
| The country has quite a good facility to prevent covid-19 | 9 | 0.57 |
| **Very poor facilities are available so far** | **1111** | **69.92** |
| *Are the people in your area/district already panic about covid-19?* |  |  |
| May be | 338 | 21.27 |
| No | 302 | 19.01 |
| **Yes** | **949** | **59.70** |
| *Are you feeling anxious/stressed/depressed/helpless thinking about the outbreak of covid-19?* |  |  |
| May be | 117 | 7.36 |
| No | 129 | 8.12 |
| **Yes** | **1343** | **84.52** |
| *If above answer is YES, then please rate your level of feelings.* |  |  |
| Extreme | 276 | 20.55 |
| **High** | **612** | **45.57** |
| Little | 66 | 4.91 |
| Moderate | 389 | 28.97 |

Table 4 shows the frequency distribution of respondents’ attitudes towards COVID-19. As the majority of the respondents (96%) are anxious about massive fatality by COVID-19 in Bangladesh – the majority (95%) of them are willing to stay at home for two weeks upon government order. The attitude towards social distancing shows mixed outcomes as ~24% of the respondents is doubtful about its efficacy. However, the actual situation - as reported in news - shows a contrary picture. The government has to deploy law enforcement to implement lock-down and social distancing [38]. The respondents have a positive attitude towards stopping business and recreational trips (96.6%), working from home (98.4%). Attitude regarding the government initiatives shows pessimism as 91.5% feel the adoption of preventive measures are inadequate and 86% feel the public officials lag in pre-emptive preparations after learning about the spread of novel coronavirus from Wuhan. Particularly, the majority (~88%) considers measures to protect healthcare professionals is inadequate as they (~88%) feel the elevated risks of COVID-19 on healthcare professionals. However, almost one third of the respondents (32%) are skeptical to consider that novel corona virus will cause pandemic in Bangladesh. Overall, in stark difference to their dissenting attitude towards the public sectors’ readiness related to the adequacy of preventive measures (91.5%) or need for pre-emptive measures (~80%) or arrangement to safeguard the healthcare providers (~88%), respondents show positive attitude to sacrifice their autonomy for the containment of contagion and towards healthcare providers in perceiving their risks and need to protect them (~88%). The attitude score and related responses clearly indicate a bleak perception towards the readiness of the government in tackling the unfolding challenges of COVID-19 management and arrested the need for more forthcoming and transparent attitude on part of the government to take opportunity of mobilizing their willingness to sacrifice for a desired outcome. Experts also opined that there was a lack of coordination towards the management of COVID-19 in the country although we had enough time to take appropriate measures [27].

On the practice side (Table 5), as the majority of the respondents (~92%) are somewhat stressed about COVID-19 outbreak in Bangladesh leading to a high to extreme feeling about the risk in 65% of them −80.5% of the claims that they avoid crowded areas and 94% do not allow children in outdoor activities and prefer using masks (~84%) while going out. In the reality of Bangladesh’s wage-dominated job sector, it is good to see that ~60% of the respondents are working from home full-time while ~14% work occasionally from home. It may not be reflective of the reality since the internet-based survey excludes responses from low-income for whom working from home is not an option. Approximately 61% of them think that awareness level among general people are little or increasing and this may explain their mixed opinion regarding COVID-19 related panic in their respective areas. As the majority of the respondents expect high possibility of spread of COVID-19 with high fatality as many believe death to be the outcome of the disease – COVID-19 may create moderate to extreme stress or anxiety among most of the respondents. Policy makers need to take heed of this in addressing the psychological aspect of the pandemic.

### Attitudes and practices in relation to knowledge and socio-socio-demographic variables

The practice of any population regarding a particular issue depends on their understanding of the issue and attitude towards it. As shown in Table 6, in all, 47.5% of the respondents belonged to the group ‘*poor knowledge with poor attitude’* and 45.6% to the group ‘*poor knowledge with poor practice*’ Among the respondents, 29.2% showed good knowledge, 35% showed good attitudes and 35.3% showed good practices. Among the people with *good knowledge*, 60% showed *poor attitude*. Interestingly, among the respondents showing *poor knowledge* scores, one-third showed *good practice* score. Results indicate the challenges of managing a pandemic on part of the government both in policy making and in implementing mitigation measures since people, even with good knowledge, may not be expected to behave accordingly. This is also reflected in respondents’ expectations of high fatality and high contagion while showing poor awareness on the reasons of contagion. Unfortunately, most of the respondents couldn’t decide the priorities of government’s action in preventing the disease while showing dissents on its preparedness. The Government of Bangladesh arranged measures and ordered for home quarantine of returnee expatriates from infected countries [24,39,42]. However, violations and even protests against government’s order is evident in many cases [37] under the excuse of inappropriate institutional quarantine facilities [52].

**Table 6.**
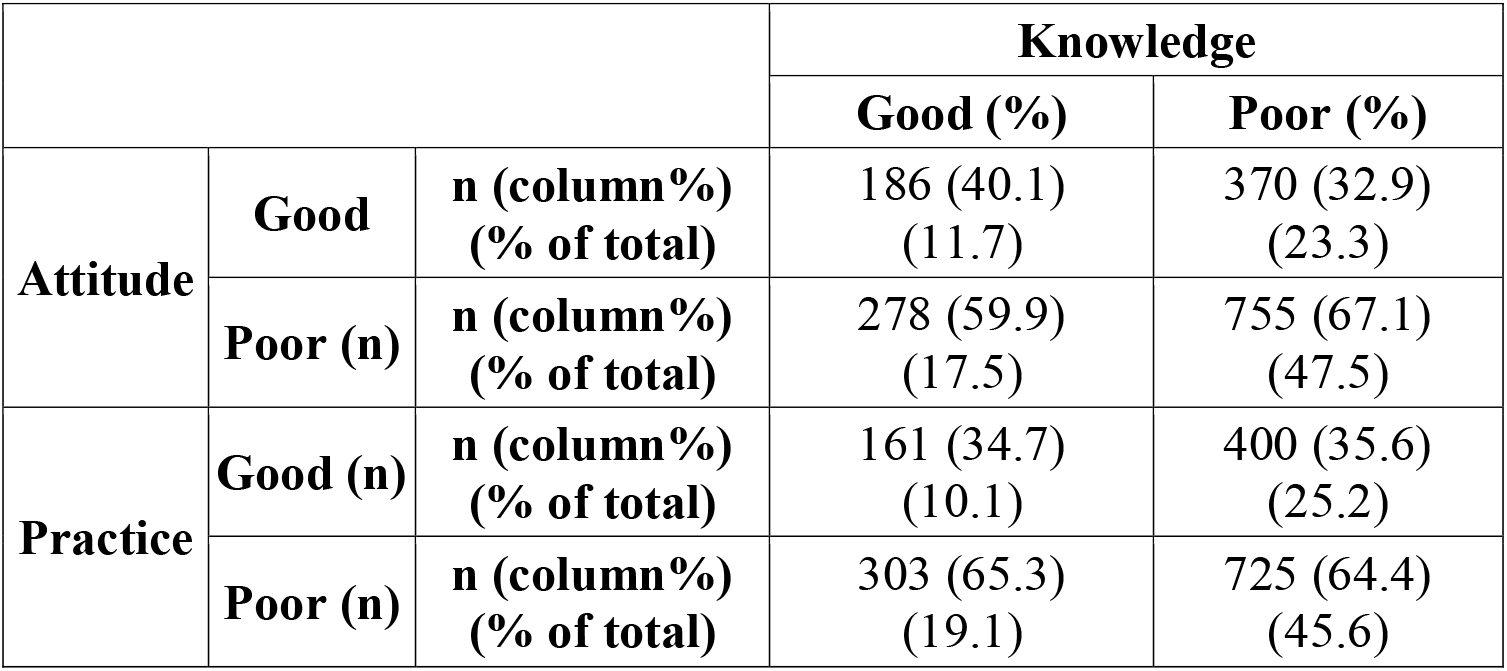
Knowledge on COVID and its relationship with practice and attitude of respondents.

|  |  |  | **Knowledge** |  |
| --- | --- | --- | --- | --- |
|  |  |  | **Good (%)** | **Poor (%)** |
| **Attitude** | **Good** | **n (column%)**<br>**(% of total)** | 186 (40.1)<br>(11.7) | 370 (32.9)<br>(23.3) |
|  | **Poor (n)** | **n (column%)**<br>**(% of total)** | 278 (59.9)<br>(17.5) | 755 (67.1)<br>(47.5) |
| **Practice** | **Good (n)** | **n (column%)**<br>**(% of total)** | 161 (34.7)<br>(10.1) | 400 (35.6)<br>(25.2) |
|  | **Poor (n)** | **n (column%)**<br>**(% of total)** | 303 (65.3)<br>(19.1) | 725 (64.4)<br>(45.6) |

Gender, age, occupation and knowledge scores show strong bearing on respondents’ opinions in both attitude and practice responses in logistic regression analysis (Table 7). It is evident that people with a better understanding of COVID-19 favour social distancing (OR 2.089, P<0.01) or working from home (OR 2.225, P<0.01). They have a better attitude towards the heightened risks of COVID-19 casualty of health care sector (OR 2.056, P<0.01) while showing a general dissatisfaction towards early response from the government on COVID-19 (OR 1.474, P<0.05) and provision of protection for healthcare workers (OR 1.47, P<0.01). This clearly indicates the need on part of the policy makers to make people educated through awareness campaigns as knowledge creates a more positive attitude and harness better practice and makes the people more capable in constructive criticism. Women are more willing than men to maintain social distances (OR 1.86, P<0.01), cancel trips (OR 2.48, P<0.05), work from home (OR 2.12, P<0.01). They also expected better pre-emptive responses from the government (OR 2.00, P<0.01) and perceived disproportionate threat of COVID-19 on healthcare workers (OR 1.57, P<0.01). Among the age groups, in reference to *‘18-25’* age group, in foregoing trips in COVID-19 *‘36-45’* years old are extremely sensible (OR 10.46, P<0.05) while the same group showed severs dissatisfaction of government’s responses (OR 0.352, P<0.01) which may be due to the inordinate involvement of this group with outdoor trips than other age groups. The response of this age group also indicates that people facing any situation directly are more likely to exhibit a more realistic response. Expectedly, in reference to government staff, professionals showed higher reluctance towards the government’s measures (OR 2.9, P<0.01). Interestingly, students (OR 0.381, P<0.01) and unemployed (OR 0.383, P<0.01) are less willing towards social distancing.

**Table 7.**
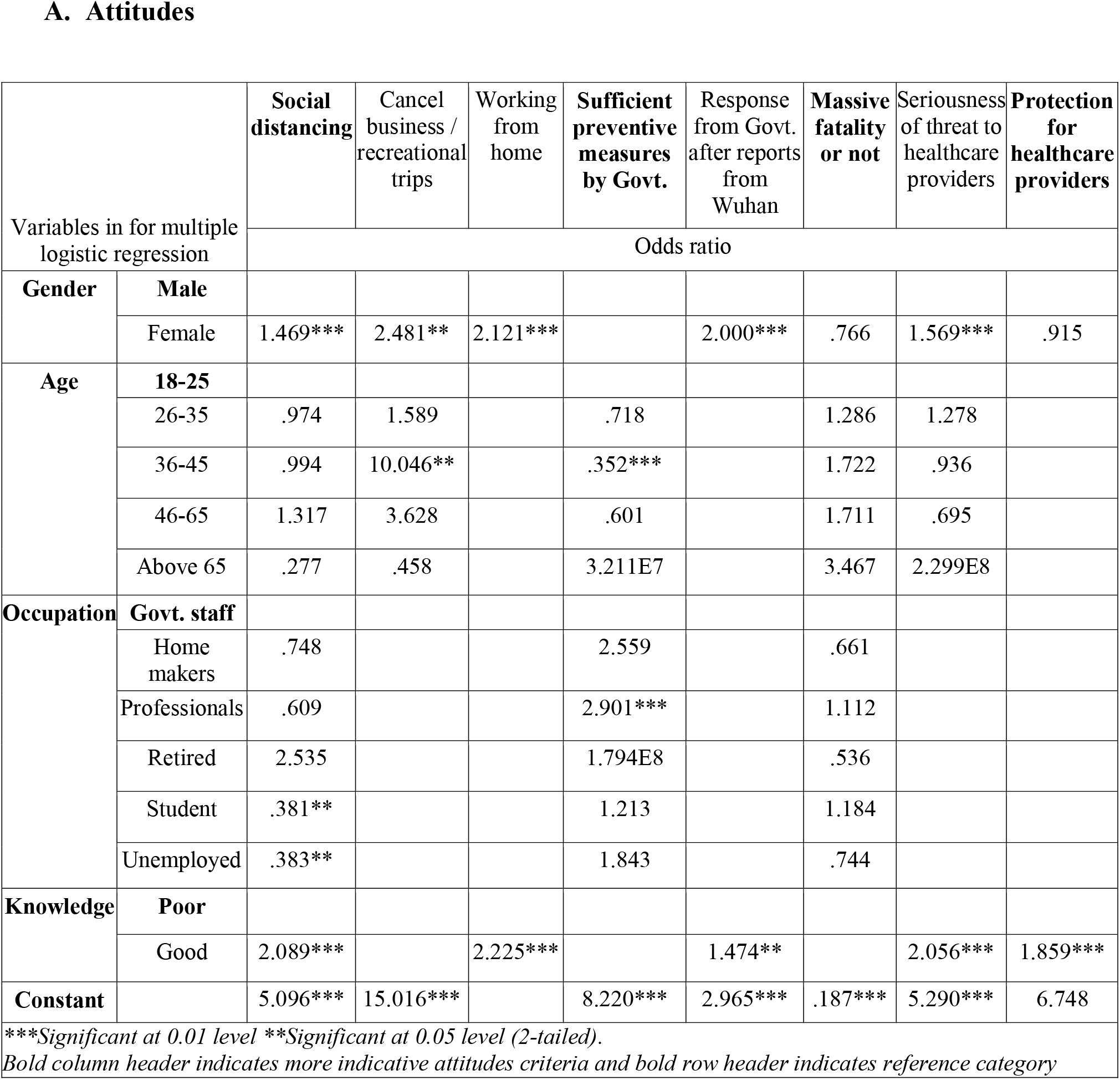
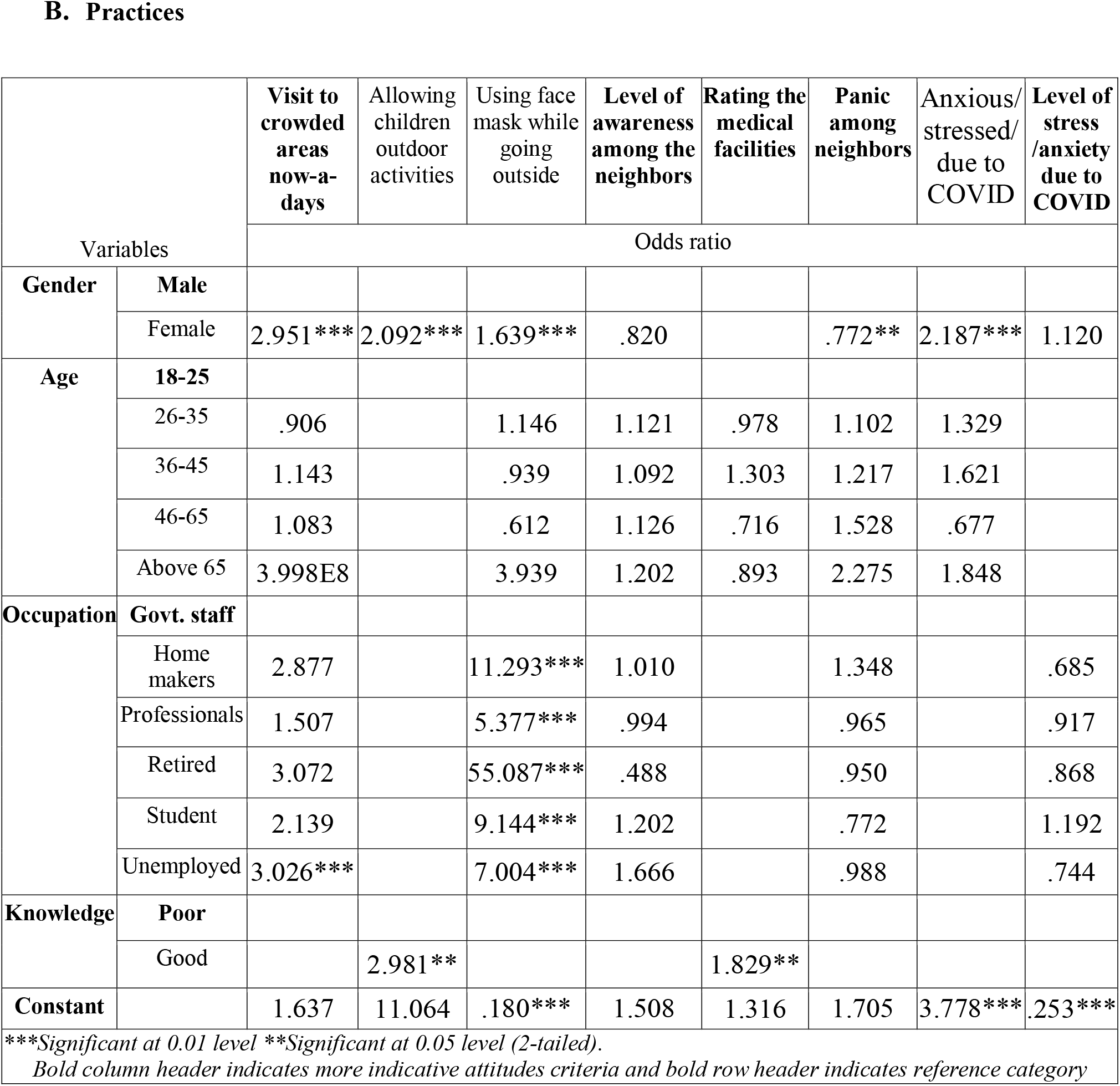
Multiple logistic regression of attitude and practice scores with socio-demographic variable and knowledge scores.

## Conclusion

COVID-19 pandemic, a disaster of magnitude unforeseen by current generations of mankind, brings the entire world into a standstill and exposed the inherent weaknesses in public health management associated with it in the developed world let alone in the developing and underdeveloped countries. Knowledge, attitude and practice of general people is crucial to the containment of this disease and in implementing mitigation measures since it arrests the participation of all to ensure social distancing, isolation, quarantine, maintaining personal and community hygiene and empathy to all who are fighting against COVID-19. An understanding of the KAP of people, hence, immensely help the policy makers and public health managers to design and implement policies and mitigation measures by providing them with insights on pertinent crucial aspects. Accordingly, this study, for the first time in Bangladesh and its peer countries, tries to evaluate the KAP of Bangladeshi people through an online survey under the ongoing social distancing requirements. Based on the valid responses from 1589 people, the study evaluates the KAP scores beside relating the scores with their socio-demographic profiles. The study concluded poor KAP among people on COVID-19 as the respective percentages of people showing poor scores in knowledge, attitude and practice are 70.8%, 65% and 64.7% with some significant bearings of socio-demographic factors on the scores. Women showed better attitude and practices despite lower knowledge score than men while retired people and homemakers indicated better attitude and practices than occupational groups. Unfortunately, KAP of people seemed unaffected by formal education level and surprisingly, the students as well as the public service professionals scored poor in attitude and practice scores. The study indicated a need for more curated awareness programmes with differential targeting and messages for different socio-demographic groups specially for students and public service professionals. An overwhelming panic was indicated as implicated by poor knowledge on fatality rate, underlying factors thereof and the general dissent on the measures hitherto adopted. Also, the study indicated poor translation of knowledge into attitude and practices, as only about 10% of the respondents showed good knowledge with good attitude and practice under a situation that requires improved attitude and practices to ensure social distancing, quarantine adoption of protective measures including wearing masks. As 99% of the respondents failed to identify the priority actions of Government in combating the disease, the Government needs to communicate more transparently to gain public confidence regarding factual information on preventive measures and its effectiveness to the people so that people don’t panic and they spontaneously follow the measures. The survey, being conducted online by and English language questionnaire, inherently included the literate persons and it can therefore be concluded that the KAP scenarios is more disappointing among lower education status. However, based on the outcome of this study, it can be claimed that routine KAP analysis can become an effective monitoring tool to measure the performance of mitigation measures in COVID-19 and beyond. And in any such application, the results of this study can be used as a baseline.

## Data Availability

Data (after analysis) are embedded in the manuscript, and raw data can be available upon request

## Acknowledgements

The team is thankful the anonymous respondents for volunteering their participation in the study.

## Supporting information

**S1 Fig. Map of Bangladesh showing the geographical distribution of respondents**.

KAP-COVID-19-Supplementary Figure.

**S1 Table. KAP questionnaire on COVID-19 for online survey across Bangladesh**.

KAP-COVID-19-Supplementary Table1.doc

**S2 Table. Pearson’s correlation coefficients for knowledge, attitude and practice**.

KAP-COVID-19-Supplementary Table2.doc

